# Energetics and Clinical Factors for the Time Required to Walk 400 Meters The Study of Muscle, Mobility and Aging (SOMMA)

**DOI:** 10.1101/2023.11.10.23298299

**Authors:** Steven R. Cummings, Li-Yung Lui, Nancy W. Glynn, Theresa Mau, Peggy M. Cawthon, Stephen B. Kritchevsky, Paul M. Coen, Bret Goodpaster, David J. Marcinek, Russell T. Hepple, Sheena Patel, Anne B. Newman

## Abstract

**Background:** Walking slows with aging often leading to mobility disability. Mitochondrial energetics has been found to influence gait speed over short distances. Additionally, walking is a complex activity but few clinical factors that may influence walk time have been studied.

**Methods:** We examined 879 participants ≥70 years and measured the time to walk 400m. We tested the hypothesis that decreased mitochondrial energetics by respirometry in muscle biopsies and magnetic resonance spectroscopy in the thigh, is associated with longer time to walk 400m. We also used cardiopulmonary exercise testing to assess the energetic costs of walking: maximum oxygen consumption (VO_2_peak) and energy cost-capacity (the ratio of VO2, at a slow speed to VO_2_peak). In addition, we tested the hypothesis that selected clinical factors would also be associated with 400m walk time.

**Results:** Lower Max OXPHOS was associated with longer walk time and the association was explained by the energetics costs of walking, leg power and weight. Additionally, a multivariate model revealed that longer walk time was also significantly associated with lower VO_2_peak, greater cost-capacity ratio, weaker leg power, heavier weight, hip and knee stiffness, peripheral neuropathy, greater perceived exertion while walking slowly, greater physical fatigability, less moderate-to-vigorous exercise, less sedentary time and anemia. Significant associations between age, sex, muscle mass, and peripheral artery disease with 400m walk time were explained by other clinical and physiologic factors.

**Conclusions:** Lower mitochondrial energetics is associated with needing more time to walk 400m. This supports the value of developing interventions to improve mitochondrial energetics. Additionally, doing more moderate-to-vigorous exercise, increasing leg power, reducing weight, treating hip and knee stiffness, and screening for and treating anemia may reduce the time required to walk 400m and reduce the risk of mobility disability.

---

Mobility, the ability to walk far enough fast enough to conduct activities of daily living, is essential for maintaining independence. The prevalence of mobility disability defined as the inability to walk ½ mile or climb a flight of steps increased from 25.9% of women and 16.7% of men aged 70-74 years reported, to 50.5% of women and 35.3% of men aged 80-84.^1^ Walking 400m is equivalent to walking 2 to 3 blocks and walking that distance too slowly may limit the ability to shop for oneself or cross intersections within the cycle of changing lights. Several studies have shown that slower 400-meter (400m) walk speed or inability to complete a 400m walk is strongly associated with progression to more advanced disability and total as well as cardiovascular mortality in older adults.^2–4^ The 6-minute walk test, a similar test that sets the time and measures distance,^5,6^ has been associated with mortality and cardiovascular events in cohort studies,^7^ and in patients with heart failure, chronic obstructive pulmonary disease, and peripheral artery disease.^8–10^

Mobility disability, requiring more than 15 minutes to walk 400m, has been the outcome for several large clinical trials.^11^ In the Lifestyle Intervention and Independence for Elders (LIFE) Study, a physical activity program reduced the incidence of major mobility disability, defined as the inability to walk 400m, by 18% from 35.5% to 30.1% after 2.6 years.^12^ The Sarcopenia and Physical fRailty IN older people: multi-componenT Treatment strategies (SPRINTT) project had interventions which reduced major mobility disability by 23%.^13^ While these findings are promising, stronger interventions are needed and might involve additional interventions on other factors that contribute to a long 400-meter walk time.^14,15^

Walking is a complex activity.^16^ Preventing decline in mobility requires a better understanding of factors that contribute to the ability to walk 400m. Multimorbidity and lack of physical activity are major correlates of mobility disability^17–19^ but do not explain the steep increase in incidence of mobility disability with age.^1,20^

Previous studies suggest that the decline of muscle mitochondrial function that occurs with aging may contribute to the decline in mobility.^21–24^ Most have used magnetic resonance spectroscopy (MRS) to estimate the capacity of mitochondria in thigh muscle to generate adenosine triphosphate (ATP).^24^ Few have also directly measured mitochondrial energetics in muscle biopsies to determine the role of muscle mitochondria in mobility.^22,25^ The energy required to walk at one’s usual pace increases with aging after the seventh decade and has been associated with slow gait speed over short distances, e.g. 6 meters.^26^

Few other potential correlates of gait speed have been studied.^16^ A few studies of gait speed over 3 to 20 m found associations that include leg extension force, height, body mass index, balance, sedentary behavior, and chronic obstructive lung and cardiovascular disease. Loss of skeletal muscle mass has been shown to play a role as well.^27^ It has been proposed that gait mechanics and stiffness of tendons around lower extremity joints might be important.^16^

We tested the hypothesis that muscle mitochondrial energetics would be an important correlate of the time it takes an individual to walk 400m. As walking 400m is a complex activity we also tested the hypothesis and systematically assessed many other factors that would contribute to the 400m walk time. We analyzed these potential correlates of the 400m walk time in the Study of Muscle, Mobility and Aging (SOMMA), a cohort study of the biological determinants of mobility.

## Methods

SOMMA is a cohort study of 879 individuals aged 70 or older who were recruited by field centers at University of Pittsburgh and Wake Forest University School of Medicine. A description of the design and methods of SOMMA has been published.^28^ All participants provided written, informed consent, and the Western IRB-Copernicus Group (WCG) Institutional Review Board approved the SOMMA (WCGIRB #20180764).

Participants must have been able to complete the 400-meter walk; those who appeared as they might not be able to complete the 400m walk at the in-person screening visit completed a short distance walk (4 meters) to ensure their walking speed was ≥0.6m/s. The 400m walk was conducted at the participant’s usual or preferred pace for 10 laps around a 40-meter course without any assistive device other than a straight cane. The total time (seconds) to walk 400m included the rest time if the participant stopped walking during the test.

To assess mitochondrial energetics, we performed high resolution respirometry on permeabilized muscle fibers from percutaneous muscle biopsies from the vastus lateralis.^22^ Methods for mitochondrial respirometry to assess the activity of mitochondrial electron transport system have been published.^25^ This study uses maximal oxidative phosphorylation (OXPHOS) supported by complex I- and II-linked carbohydrates (Max OXPHOS) as a measure of *ex vivo* muscle mitochondrial energetics. In addition, we used ^31^Phosphorous magnetic resonance spectroscopy (^31^P MRS) to measure of *in vivo* muscle mitochondrial energetics. It measures the rate of phosphocreatine (PCr) recovery after an acute bout of knee extensor exercise to estimate the maximal production of adenosine triphosphate (ATP_max_).^29^

We defined several domains of factors, besides mitochondrial energetics, that may contribute to the time required to complete the 400m walk. The domains were cardiorespiratory fitness and energetic cost of walking, body stature and composition, muscle mass, strength and power, blood flow, oxygen delivery, cardiopulmonary disease, perceived fatigability and exertion, depressive symptoms, cognitive performance, neurologic conditions, joint symptoms, usual physical activity, vision, general health and medical conditions, health habits, and socioeconomic status. For each domain, we measured candidate variables that reflect the effect of the domains on time to walk 400m.^28^

To assess the energetic costs of walking, participants completed a three-stage cardiopulmonary treadmill exercise test (CPET) using a symptom-limited modified Balke or manual protocol during which the volume of oxygen (VO_2_) uptake was measured.^30^ Participants began the VO_2_peak test at preferred walking speed as the treadmill speed (0.5 mph) and grade (2.5%) were incrementally increased until participants reach a respiratory exchange ratio (RER) ≥1.05 and self-reported Borg Rating of Perceived Exertion (RPE) was ≥17.^31^ VO_2_peak (mL/min) was defined as the highest 30-second average oxygen consumption over the course of the test. The energy cost of walking was calculated as the average VO_2_ mL/min while walking at a steady slow speed: 1.5 mph for 5 min at zero grade. Energy cost-to-capacity ratio is the proportion of VO_2_ peak required to walk at the steady 1.5 mph pace (the ratio of VO_2_ mL/min at a steady pace divided by VO_2_peak mL/min.^32^

Weight was measured using a balance beam or digital scales. Leg extension strength was assessed by a one repetition max (1RM) on a Keiser Air 420 exercise machine, and peak power was measured across the 40-70% range of leg extension.^33^ Whole body skeletal muscle mass was measured by D_3_-creatine dilution (D_3_Cr).^34,35^ A whole body magnetic resonance (MR) scan included fat-free thigh muscle volume.^36^

To assess impaired delivery of oxygen to leg muscles, systolic blood pressure was measured by Doppler in the arm (brachial artery) and ankle (posterior tibial artery) on both sides and peripheral artery disease was classified as present if the ankle to brachial artery ratio or index (ABI) was less than 1.0 in either leg.^37,38^ Anemia was defined as hemoglobin <13.5% for men and <12% for women. We measured perceived fatigability by the Pittsburgh Fatigability Scale (PFS, scale 0-50, higher = greater fatigability) and Borg RPE Scale (RPE Fatigability, scale 6-20, higher exertion = greater fatigability) at the end of the 5 min walk.^39,40^

We assessed peripheral sensory neuropathy by reduced touch sensation by the ability to feel fine filaments pressed against the great toe.^41^ Joint stiffness was assessed as self-report of stiffness in the hip or knee that made it difficult to walk often or always in the past 7 days.

Physical activity was assessed using a wrist-worn ActiGraph GT9X for 7-full days and we assessed the time spent in moderate-to-vigorous physical activity (MVPA) (minutes/day).^42,43^ Participants concurrently wore an activPal device on the thigh to characterize sedentary time (minutes/day) and total daily step count. General health status and medical history was assessed by self-reported diagnosis of common medical conditions. Methods for other examinations are described in the Supplement.

## Statistical Methods

Linear regression was used in age-adjusted analyses to analyze the association of each variable with 400m walk time. We then used a series of forward stepwise regression models using a P-value of 0.1, to select the final multivariate model to identify the correlates of 400m walk time. Lastly, we used multiple imputation by fully conditional specification method to account for missing data (Supplemental Table 1). The results are expressed in seconds of walk time per SD or unit of each potential correlate or the equivalent beta coefficients.

We expected that many factors, such as mitochondrial energetics, would influence 400m walk time by their effect on other assessments, such as the energy cost of walking and may, therefore, not appear in the multivariable model. Thus, we systematically analyzed how the association of muscle mitochondrial energetics (Max OXPHOS) with 400m walk time was explained by VO_2_peak, the ratio of energy cost-capacity, and leg power by adding them individually and then together to the model between Max OXPHOS and 400m walk time.

Based on expert judgement and prior evidence about factors that influence walking speed, we analyzed other variables that were associated with age-adjusted 400m walk in bivariate analyses but were not included in the final stepwise model: age, sex, muscle mass and peripheral artery disease with walk time. As with the analysis of Max OXPHOS, we started with the association of that factor with 400m walk time, then adjusted for variables from the multivariable model that may at least partially account for the association until the association was no longer statistically significant.

## Results

Characteristics of the 879 participants are described in Table 1. Participants had no contraindications to MR, CT scans, or muscle biopsy and, at a screening visit, were able to walk at least 0.6 m/s. The participants were initially contacted and screened for eligibility by phone.

**Table 1.** Selected characteristics of the SOMMA cohort.

| Characteristic* |  |
| --- | --- |
| Sex | N (%) |
| Men (%) | 359 (40.8) |
| Women (%) | 520 (59.2) |
| Race | N (%) |
| White (%) | 745 (84.8) |
| Black (%) | 116 (13.2) |
| American Indian/Alaskan native (%) | 2 (0.2) |
| Asian (%) | 6 (0.7) |
| Multiracial/Unknown (%) | 10 (1.0) |
| Hispanic/Latino | 9 (1.1) |
| Age, years (SD) | 76.3 (5.0) |
| Age groups | N (%) |
| 70-74 | 402 (45.7) |
| 75-79 | 273 (31.1) |
| 80-84 | 134 (15.2) |
| 85+ | 70 (8.0) |
| 400m walk time (min); mean (SD) | 6.6 (1.2) |
| 400m walk time groups (min) | N (%) |
| <6 | 308 (35.0) |
| 6 –< 8 | 473 (53.8) |
| 8 – 15 | 98 (11.2) |
| Weight (kg) | Mean (SD) |
| Men | 84.8 (13.8) |
| Women | 70.1 (13.0) |
| Body mass index (kg/m<sup>2</sup>); mean (SD) | 27.6 (4.6) |
| Peak leg power (Watts) | Mean (SD) |
| Men | 479.8 (161.4) |
| Women | 271.0 (84.8) |
| Time in moderate to vigorous activity (min/d); mean (SD) | 186.5 (86.1) |
| Sedentary time excluding in-bed time (min/d); mean (SD) | 619.1 (113.1) |
| MoCA Total score; mean (SD) | 24.8 (2.9) |
| Anemia, N (%) | 142 (16.4) |
| History of chronic obstructive pulmonary disease, N (%) | 115 (13.1) |
| History of heart failure, N (%) | 6 (0.7) |
| History of stroke, N (%) | 21 (2.4) |
| History of diabetes, N (%) | 131 (15.0) |
| History of a diagnosis of depression, N (%) | 161 (18.4) |
| Current smoker, N (%) | 25 (2.9) |
| Number of drinks in the past 12-month, N (%) | 2.8 (4.5) |
| Ankle-arm blood pressure index (Lowest of Left and Right) < 1.0, N (%) | 97 (12.0) |
| Peripheral neuropathy by monofilament test, N (%) | 46 (5.3) |
| Often/always hip or knee stiffness, N (%) | 14 (1.6) |
| Pittsburgh Fatigability Scale Physical score, mean (SD) | 15.8 (8.7) |
| Rating of Perceived Exertion Fatigability at end of the steady speed test, mean (SD) | 8.4 (1.9) |
| VO<sub>2</sub>peak (volume of oxygen consumption) (mL/min) | mean (SD) |
| Men | 1865.6 (410.7) |
| Women | 1287.0 (248.9) |
| Energy cost-capacity ratio, mean (SD) | 48.8 (12.6) |
| Maximal mitochondrial oxidative phosphorylation (pmol/(s*mg), mean (SD) | 59.7 (18.5) |

\*N (%) for categorical variables and mean (SD) for continuous variable

Of those eligible for and attended a screening 80% were enrolled in the study (Supplemental Figure 1). Details of process and results of screening have been published.^28^

About half of the cohort was between 70 and 74 years of age. The mean 400-meter walk time was 6 minutes, 36 seconds.

There were weak or no correlations between the measurements except for an expected correlation of Pearson’s r > 0.5 between measurements of body mass, strength, power and weight with VO_2_peak (Supplemental Figure 2). Most of the 53 variables that were significantly associated with walk time remained so after adjusting for age (Table 2).

**Table 2.** List of candidate factors and their bivariate associations with 400m walk time. The domain category is in bold.

| **Domain** and measurement | Age-adjusted<br>beta coefficient<br>(95% CI) per SD | Age-adjusted<br>p-value |
| --- | --- | --- |
| Age* (years) | -- | -- |
| Sex* | 23.0<br>(13.8, 32.2) | <0.0001 |
| Race* (non-Hispanic White) | 43.3<br>(30.5, 56.0) | <0.0001 |
| Education (College graduate or post college) | 17.2<br>(7.8, 26.7) | 0.0004 |
| **Leg muscle energetics** |  |  |
| Complex I and II supported Maximal OXPHOS (pmol/(s*mg)) | 22.1<br>(17.5, 26.8) | <0.0001 |
| ATP<sub>max</sub> (mM/sec) | 12.4<br>(7.7, 17.2) | <0.0001 |
| **Cardiorespiratory fitness and energetics of walking** |  |  |
| VO<sub>2</sub>peak (volume of oxygen consumption) (mL/min) | 19.1<br>(14.4, 23.8) | <0.0001 |
| Energy Cost-Capacity Ratio (%) | 37.7<br>(33.5, 41.9) | <0.0001 |
| **Body stature and composition** |  |  |
| Height (m) | 10.8<br>(6.3, 15.4) | <0.0001 |
| Weight (kg) | 14.6<br>(2.3, 10.1) | <0.0001 |
| Body mass index (kg/m<sup>2</sup>) | 25.8<br>(21.5, 30.1) | <0.0001 |
| Mean Quadriceps Muscle Fat Infiltration (%) | 27.3<br>(22.9, 31.6) | <0.0001 |
| Total Adipose Tissue (vol, L) | 24.5<br>(20.1, 28.9) | <0.0001 |
| **Muscle mass and strength** |  |  |
| D<sub>3</sub>Cr Muscle mass (kg) | 6.9<br>(2.0, 11.7) | 0.0055 |
| Total Thigh Fat Free Muscle (volume, L) | 10.0<br>(5.2, 14.7) | <0.0001 |
| Leg Power Test 1RM (kg) | 17.3<br>(12.7, 21.9) | <0.0001 |
| Peak Leg Power (Watts) from 40-70% of 1RM | 18.9<br>(14.2, 23.7) | <0.0001 |
| **Blood flow, oxygen delivery, cardiopulmonary disease** |  |  |
| Ankle-Arm Blood Pressure (Lowest of Left and Right) < 1.0 | 34.3<br>(19.7, 48.9) | <0.0001 |
| Anemia, (Hemoglobin (g/dL) <13.5 M, <12.0 F) | 29.1<br>(16.7, 41.5) | <0.0001 |
| Heart Failure | 3.9<br>(-51.6, 59.4) | 0.8903 |
| Systolic Blood Pressure (mmHg) | 2.9<br>(-1.7, 7.4) | 0.2201 |
| Diastolic Blood Pressure (mmHg) | 4.5<br>(-0.1, 9.1) | 0.0547 |
| Chronic Obstructive Pulmonary Disease* | 15.1<br>(1.6, 28.6) | 0.0291 |
| **Perceived fatigability and exertion** |  |  |
| Pittsburgh Fatigability Scale Physical score, 0-50<br>(higher=greater fatigability) | 24.3<br>(20.0, 28.7) | <0.0001 |
| Pittsburgh Fatigability Scale Mental score, 0-50 (higher=greater<br>fatigability) | 16.1<br>(11.7, 20.6) | <0.0001 |
| Ratings of Perceived Exertion (RPE) Fatigability at end of slow<br>walking speed test, (6-20; higher=greater exertion) | 27.3<br>(23.0, 31.6) | <0.0001 |
| Ratings of Perceived Exertion (RPE) Fatigability at end of 400M<br>walking test, (6-20 (higher=greater exertion) | 21.9<br>(17.5, 26.2) | <0.0001 |
| **Depressive symptoms** |  |  |
| Center of Epidemiologic Studies Depression Scale (CESD-10)<br>(0-30) | 3.5<br>(2.2, 4.8) | <0.0001 |
| Depression, self-report* | 22.5<br>(10.8, 34.3) | 0.0002 |
| **Cognitive performance** |  |  |
| Digit Symbol Substitution Test (number of correct boxes) | 19.0<br>(14.4, 23.6) | <0.0001 |
| Montreal Cognitive Assessment (MoCA) Total score (0-30) | 7.5<br>(2.8, 12.2) | 0.0017 |
| Trails B: Total time (0-300 sec) | 16.2<br>(11.5, 20.8) | <0.0001 |
| **Neurologic conditions** |  |  |
| Monofilament test result - normal | 17.7<br>(-2.9, 38.2) | 0.0924 |
| Experienced numbness, loss of sensation - Intensity | 27.4<br>(1.5, 53.2) | 0.0385 |
| History of stroke* | 1.8<br>(-2.8, 6.4) | 0.4447 |
| **Joint symptoms\*** |  |  |
| Any pain in left or right hip or knee | 21.6<br>(11.7, 31.5) | <0.0001 |
| Any back pain after 400m walk (Pain level 1-10=yes, 0=no) | 39.0<br>(23.1, 55.0) | <0.0001 |
| Moderate/Severe/Extreme hip or knee stiffness today | 46.4<br>(31.1, 61.7) | <0.0001 |
| In the past 7 days, often/always joints stiffness | 86.4<br>(50.4, 122.4) | <0.0001 |
| **Usual physical activity** |  |  |
| activPAL: Mean of Total sedentary time, excluding in-bed time<br>(min/day) | 5.4<br>(0.3, 10.5) | 0.0376 |
| activPAL: Mean of Total daily step count | 23.0<br>(18.1, 27.9) | <0.0001 |
| ActiGraph: Mean of Sum of total activity counts/min over 24hrs | 14.6<br>(10.0, 19.2) | <0.0001 |
| ActiGraph: Mean of Time spent in Moderate-to-vigorous physical activities (min/day) | 16.2<br>(11.6, 20.8) | <0.0001 |
| **Vision** |  |  |
| Pelli Robson Vision: Log contrast sensitivity | 5.4<br>(0.8, 10.1) | 0.0229 |
| Bailey Lovie Vision: Acuity 20/50 or worse | 6.7<br>(-3.8, 17.2) | 0.2136 |
| **General health and medical conditions** |  |  |
| Good/Very Good/Excellent* | 39.3<br>(18.5, 60.1) | 0.0002 |
| Diabetes* | 22.3<br>(9.6, 35.1) | 0.0006 |
| HbA1c ≥6.5% | 30.6<br>(15.8, 45.4) | 0.0001 |
| Red cell distribution width (%) | 12.1<br>(7.5, 16.7) | <0.0001 |
| **Health habits** |  |  |
| Cigarette smoking status* | 45.7<br>(18.4, 73.0) | 0.0011 |
| Average drinks per week in past 12 months* | 9.6<br>(5.1, 14.2) | <0.0001 |
| Number of prescription medications in the past 30 days* | 15.0<br>(10.6, 19.5) | <0.0001 |
| **Socioeconomic status** |  |  |
| Financial Accounts/Wealth* (0-8) | 16.5<br>(12.0, 21.0) | <0.0001 |

Beta coefficients represent seconds of walk time.
Notes. \*self-reported history.
Abbreviations: D<sub>3</sub>Cr: d3-creatine dilution; 1RM, 1-repetition max; ATP<sub>max</sub>, maximal adenosine triphosphate; OXPHOS, oxidative phosphorylation; HbA1c, hemoglobin A1C.

In bivariate analyses, maximal muscle mitochondrial respiration during ATP production (Max OXPHOS) was strongly associated with 400m walk time: an additional 22.1 s (17.5, 26.8) for each SD decrement. The association between max OXPHOS, with 400m walk time was completely attenuated by adjustments for the energy cost-capacity ratio, leg power, cardiorespiratory fitness (VO_2_peak) and weight (Table 3). This accounts for the fact that Max OXPHOS, did not appear in the multivariate model that included these variables. The association between ATP_max_ and 400m walk time was no longer significant after adjustment for a higher energy cost-capacity ratio and heavier weight. Furthermore, the association of *in vivo* ATP_max_ by ^31^P MRS with walk time (beta coefficient=12.4, P<0.001) was completely attenuated by adjustment for Max OXPHOS measured *ex vivo* from the muscle biopsy (adjusted beta coefficient = 3.6, p = 0.21). Therefore, hereafter, ‘mitochondrial energetics’ refers to Max OXPHOS.

In the multivariate model a higher ratio of VO_2_ at the slow walking speed to the VO_2_peak (cost-capacity ratio) was associated with nearly 20 seconds longer per SD of the measurement, to compete the 400m walk (Table 4). In addition, a lower VO_2_peak indicated a 9 second longer time, per SD, to walk 400m. Weaker leg power was associated with about 17 seconds slower time for each standard deviation (SD) decrement. Despite the greater leg power of heavier individuals, greater weight was associated with longer walk time—adding about 22 seconds for each 15 kg greater weight. Self-report of frequent or continuous stiffness in the hips or knees was associated with about 54 seconds longer time to walk 400m. Peripheral neuropathy by the monofilament test added 20 seconds to the walk time.

**Table 4.** Multivariable model of measurements associated with a slower time to complete a 400m walk.

| Measurement | Unit or S.D. | Slower time (s)<br>(95% CI) per unit<br>or per SD* | p-value |
| --- | --- | --- | --- |
| Higher energy cost-capacity ratio | 13.1% | 19.9<br>(13.8, 26.0) | <.001 |
| Lower VO<sub>2</sub>peak | 445.1 mL/min | 9.3<br>(2.2, 16.4) | 0.011 |
| Weaker peak leg power | 161.0 watts | 17.2<br>(11.9, 22.6) | <.001 |
| Greater weight | 15 kg | 21.5<br>(16.0, 27.0) | <.001 |
| Frequent or constant hip or knee joint stiffness | 1 (yes) | 53.7<br>(24.6, 82.8) | <.001 |
| Peripheral neuropathy by monofilaments | 1 (abnormal) | 20.2<br>(3.9, 36.4) | 0.015 |
| Greater perceived exertion at end of steady state walking test† | 2 units | 10.0<br>(5.3, 14.8) | <.001 |
| Higher Pittsburgh Fatigability Scale Physical score | 8.7 units | 6.0<br>(1.9, 10.1) | 0.004 |
| Less time in moderate-to-vigorous activity min/d | 88.5 minutes | 8.7<br>(4.2, 13.2) | <.001 |
| Less sedentary time minutes/d | 127.3 min | 9.2<br>(3.8, 14.5) | <.001 |
| Anemia | 1 (anemia) | 15.6<br>(5.9, 25.3) | 0.002 |

\* Equivalent to the beta coefficient
† On the 6- to 20-point Borg scale

Greater perceived physical fatigability was also associated with slower time to walk 400m by 6.0 (1.9, 10.1) seconds. Every 2-unit increase on the 6–20-point Borg scale, during the last 3 of 5 minutes of walking at a slow speed was associated with 10 seconds longer to complete the walk. Less time spent in objectively measured habitual moderate to vigorous physical activity was associated with a 9-seconds longer walk time for each decrement of SD. Less time spent in a sedentary mode was also associated with longer time to complete the 400-meter walk in bivariate analyses. Anemia was associated with a longer time to complete the walk by 16 seconds.

Several factors that may be important for the 400-meter walk time and were significant in age-adjusted associations did not appear in the multivariate model. This suggested that their associations with walk time were accounted for by other measurements in multivariate model. Besides Max OXPHOS and ATPmax, we examined the factors that explained the associations for age, sex, muscle mass, and peripheral vascular disease (Table 3). The association of older age with longer walk time was explained by weaker leg power, a higher energy cost-to-capacity ratio, less time spent in moderate to vigorous physical activity, and the association of greater weight with walk time. The longer walk time in women than men was fully explained by weaker leg power and the association of greater weight with slower walk time. Lower muscle mass by the D_3_Cr assay has been associated with slower walking speed.^27^ In this analysis, associations of lower muscle mass by D_3_Cr or by thigh fat-free muscle volume with longer walk time were fully attenuated by adjustment for heavier weight, weaker leg power and lower VO_2_peak. In accord with previous research, peripheral vascular disease assessed by an ankle-arm blood pressure ratio <1.0, was associated with longer walking time;^44^ we found that the association was completely attenuated by weight, weaker leg power, lower VO_2_peak, higher ratio of energy cost-to-capacity, and anemia.

## Discussion

The results of this study support the hypothesis that, in older adults, decreased mitochondrial energetics is associated with a longer time required to walk 400m. Walking 400m is a complex activity, and the results also support the hypothesis that several clinical and physiologic factors, some that can be modified, also influence 400m walk time.

A few previous studies have reported correlations between muscle mitochondrial energetics by high resolution respirometry in muscle biopsies with 400m walk speed.^21–23,45^ Others have reported correlations between ATP_max_ by ^31^P MRS and slower walking speed.^23,46,47^ Our results suggest that the direct assessment of mitochondrial energetics in muscle biopsies is more strongly associated with 400m walk time than is in vivo ATP_max_. Studies have also observed that increasing acquired mitochondrial DNA mutations is associated with slower walking speed.^48^ These studies using several approaches to assess the function and integrity of mitochondria in muscle lend support to our finding that muscle mitochondrial energetics is strongly associated with the time needed to walk 400m. Thus, ongoing development of treatments that preserve or enhance mitochondrial energetics may improve mobility and reduce the risk of disability.

The influence of mitochondrial energetics (Max OXPHOS) on 400-meter walk time was explained by leg power, the ratio of energy cost-to-capacity, and VO_2_peak along with weight. This is consistent with our previous findings that mitochondrial energetics in leg skeletal muscle is associated with VO_2_peak, and with leg power.^25^ Our results confirm that, as the energy required to walk slowly approaches the capacity to generate energy, the time required to walk 400m increases.

Several other factors also influenced walk time. Weaker leg power, which partly reflects lower mitochondrial energetics, also substantially increased the time required for the 400m walk. Greater weight was associated with greater leg power, but this was countered by the greater energy required to move a heavier body, suggesting that weight loss may reduce 400m walk time. Hip and knee stiffness added nearly one minute to the time required to walk 400m. Depending on the underlying cause, these symptoms might be improved by non-steroidal anti-inflammatory drugs or physical therapy. We discovered that reduced touch sensation— measured by monofilament testing, prolonged average walking speed by about 20 seconds, suggesting that sensation is important for walking. As expected, a reduced capacity to generate energy and greater energy cost of walking resulted in a greater sense of exertion from walking slowly, as measured by the Borg Scale of perceived exertion during walking. The association of higher levels of physical fatigability during usual activity by the Pittsburgh scale indicates greater whole-body vulnerability to fatigability during standardized activities and might also reflect lower mitochondrial energetics during usual activities. It has also been shown that greater fatigability predicts greater risk of functional decline and mortality.^49,50^

The association between habitual moderate-to-vigorous physical activities and 400m walk time supports promotion of regular moderate an vigorous physical activity to reduce the risk of mobility and maintain independence with aging.^49^ This result is consistent with the results of LIFE Study randomized trial that found that a structured, moderate-intensity physical activity program reduced the risk of major mobility disability defined as an inability to walk 400m within 15 minutes.^12^ The association between less sedentary time and longer walk time was counterintuitive with no apparent reason why more sedentary time in usual activities would have a beneficial effect on time to walk 400m.

Anemia was associated with about 15 seconds longer time, presumably by limiting the supply of oxygen to muscle indicating that treatment of anemia might improve mobility. This suggests that screening for and treating anemia might reduce the time required to walk 400m and forestall the development of mobility disability.^44^

It appeared that needing more time to walk 400m with aging may be at least partly attributable to decreasing leg power, a greater energy cost-to-capacity ratio, spending less time spent in at least moderate physical activity. This suggests that decline in walking speed with aging might be at least partly ameliorated by increasing physical activity and forms of exercise that improve leg power. On average, women required more time to walk 400m and this was partly attributable to lower leg power. We found that an association between lower muscle mass and longer 400-meter walk time was explained by weaker leg power and lower VO_2_peak. This suggests that increasing muscle mass might reduce the time required to walk 400m and reduce the risk of mobility disability.^27^ Peripheral artery disease also reduces blood and oxygen supply to the legs, and we found that it was associated with longer 400-meter walk time that was accounted for by its effects on reduced VO_2_peak and by weaker leg power.

This study has limitations. It is cross-sectional, based on baseline data from SOMMA. Longitudinal data would enable associations of change in some factors with change in time to walk 400m, and follow-up to identify predictors of mobility disability. However, this cross-sectional study has the advantage of associating the immediate concomitant influence of energetics and clinical factors with the present time to walk 400-meters.

SOMMA did not assess the central nervous system control of walking, the influence of impaired balance, nor the mechanics of walking.^16,52^ A small proportion of the population walked slowly, 0.6-0.8 m/s and individuals with very slow gait, <0.6 m/s, were not included. The prevalence of some conditions, such as stroke, heart disease, and severe peripheral artery disease, may have been too low to detect their impacts. While the racial composition of the study reflects that of the communities surrounding the field centers, there was insufficient power to study potential racial differences in correlates of walk time.

This study has several strengths. It is the first to use direct measurement of skeletal muscle mitochondrial energetics from biopsies and energetics of walking by cardiopulmonary exercise testing to show that the effects of mitochondrial energetics are substantially mediated by the energetics of walking and leg power. Our endpoint, the time to walk 400-meters, is important to many activities of daily living whereas previous studies assessed gait speed over shorter distances. Furthermore, SOMMA assesses a wide array of potential causes of a long 400-meter walk time in a large population that enabled this analysis to identify several independent and modifiable predictors of walk time.

## Conclusion

The time older individuals require to walk 400m is associated with muscle mitochondrial energetics. Aerobic exercise improves mitochondrial energetics and the ongoing development of novel therapies that target mitochondria might yield treatments that improve mitochondrial energetics and delay mobility disability.^53^ In addition, other factors that may be amenable to interventions also influence 400m walk time. Our results suggest that, for older patients, maintaining habitual moderate-to-vigorous exercise and promoting exercise that improves leg power, reducing weight, treatment of lower hip and knee stiffness, and screening for and treatment of anemia may reduce the time they require to walk 400m and might thereby reduce their risk of developing mobility disability.

## Data Availability

https://sommaonline.ucsf.edu/

## Acknowledgements – potential conflicts of interest

Dr. Cummings: consultant to BioAge

Dr. Cawthon: consultant to BioAge and consultant to Myocorp and owns stock in Myocorp

LL: none

NG: none

PMC: consultant to BioAge and consultant to Myocorp and owns stock in Myocorp

SBK: none

PC: none

BG: none

DJM: none

RTH: none

SP: none

ABN: none

## Author contributions

|  |  |
| --- | --- |
| Steven R. Cummings | Conceived the analysis plan, wrote 1<sup>st</sup> draft, made rounds of revisions, and wrote the final version |
| Li-Yung Lui | Did most of the statistical analyses, wrote the statistical analysis section and made edits and revisions to the text |
| Nancy W. Glynn | Made revisions on several rounds of review |
| Theresa Mau | Provided references and generated the reference list and made revisions and edits on the last 3 drafts |
| Peggy M. Cawthon | Wrote text on clinical examinations, muscle mass measurements and supplemental methods |
| Stephen B. Kritchevsky | Made revisions to early versions of the paper and oversaw the internal SOMMA review and approval of the paper |
| Paul M. Coen | Wrote sections describing muscle respirometry |
| Bret Goodpaster | Revised and approved sections regarding cardiopulmonary exercise testing |
| David J. Marcinek | Directed 31PMRS scanning and reviewed and edited the section on mitochondrial energetics |
| Russell T. Hepple | Directs SOMMA's muscle biology core. Made revisions on an early draft and reviewed and approved the final version |
| Sheena Patel | Conducted the first rounds of statistical analyses. Reviewed and approved the final version of the statistical analysis section |
| Anne B. Newman | Contributed potential predictors of 400-walk time. Wrote the 1<sup>st</sup> draft of the Introduction and then parts of the Discussion. Made revisions on the last 3 versions of the manuscript |

## Supplement: Assessment methods

Digit symbol substitution test (DSST) is a cognitive function measure of processing speed: in this pencil and paper test, the participant associates a series of symbols with corresponding numbers. Global cognitive function was by the Montreal Cognitive Assessment (MoCA) tool,^1^ and executive function by the Trails B test.^2^ Perceived physical and mental fatigability was captured using the validated 10-item Pittsburgh Fatigability Scale (PFS, 0-50, higher score equals greater fatigability).^3,4^ Body stature and composition were assessed stadiometers (height), digital scales (weight), whole body MRI analyzed by AMRA (total thigh muscle volume; muscle fat infiltration; abdominal adiposity as the sum of abdominal subcutaneous and visceral fat volume),^5^ D_3_Cr muscle mass (by d3-creatine dilution),^6,7^ and body mass index was calculated as weight (kg) divided by height (m^2^). Reduced touch sensation by the ability to feel fine filaments pressed against the great toe was used to assess peripheral neuropathy.^8^ Contrast sensitivity and visual acuity were also assessed.^9,10^ General health was measured by self-reported health and analyzed as good/very good/excellent vs. lower rating. Self-reported health habits included smoking and alcohol use. Participants were instructed to bring in all prescription medications used in the past 30 days; clinic staff recoded the number brought into clinic. Participants reported the number of financial accounts they had as a marker of wealth. Ankle-arm blood pressure was measured to assess peripheral atrial disease.^11^ Diastolic and systolic blood pressures were measured. Numbness was considered present if intensity of “numbness, loss of sensation, or a ‘dead’ feeling like an anesthetic, without prickling, in your feet or leg below the knees” in the past twenty-four hours was moderate or severe. Pain in the hip and knee was assessed using the Brief Pain Inventory administered on the day of (and before) the 400m walk assessment. A complete blood count was used to determine HbA1c and red cell distribution width.

---

**Supplemental Figure S1.**
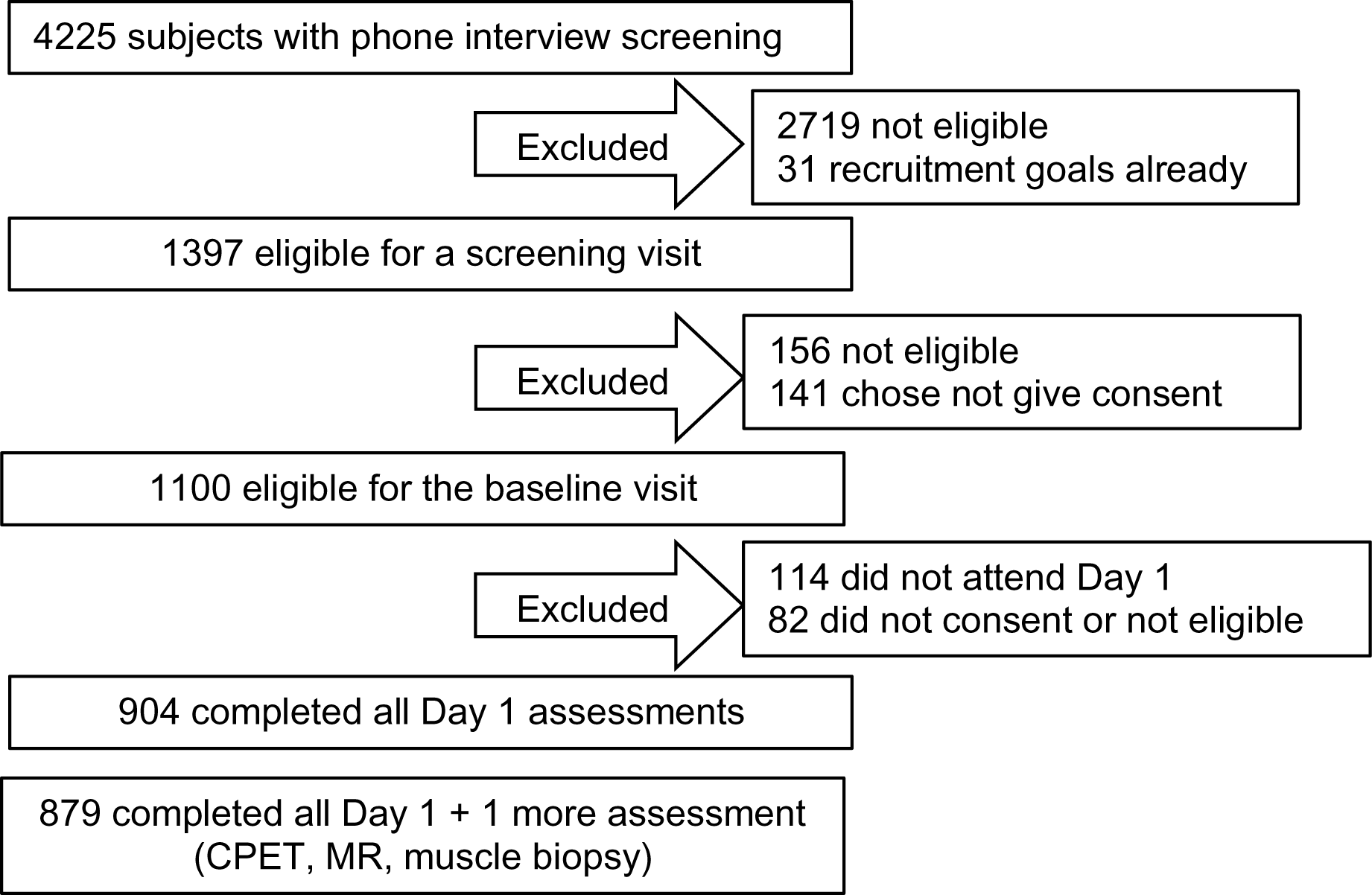
Flow chart of the enrollment process for SOMMA.

**Supplemental Figure S2.**
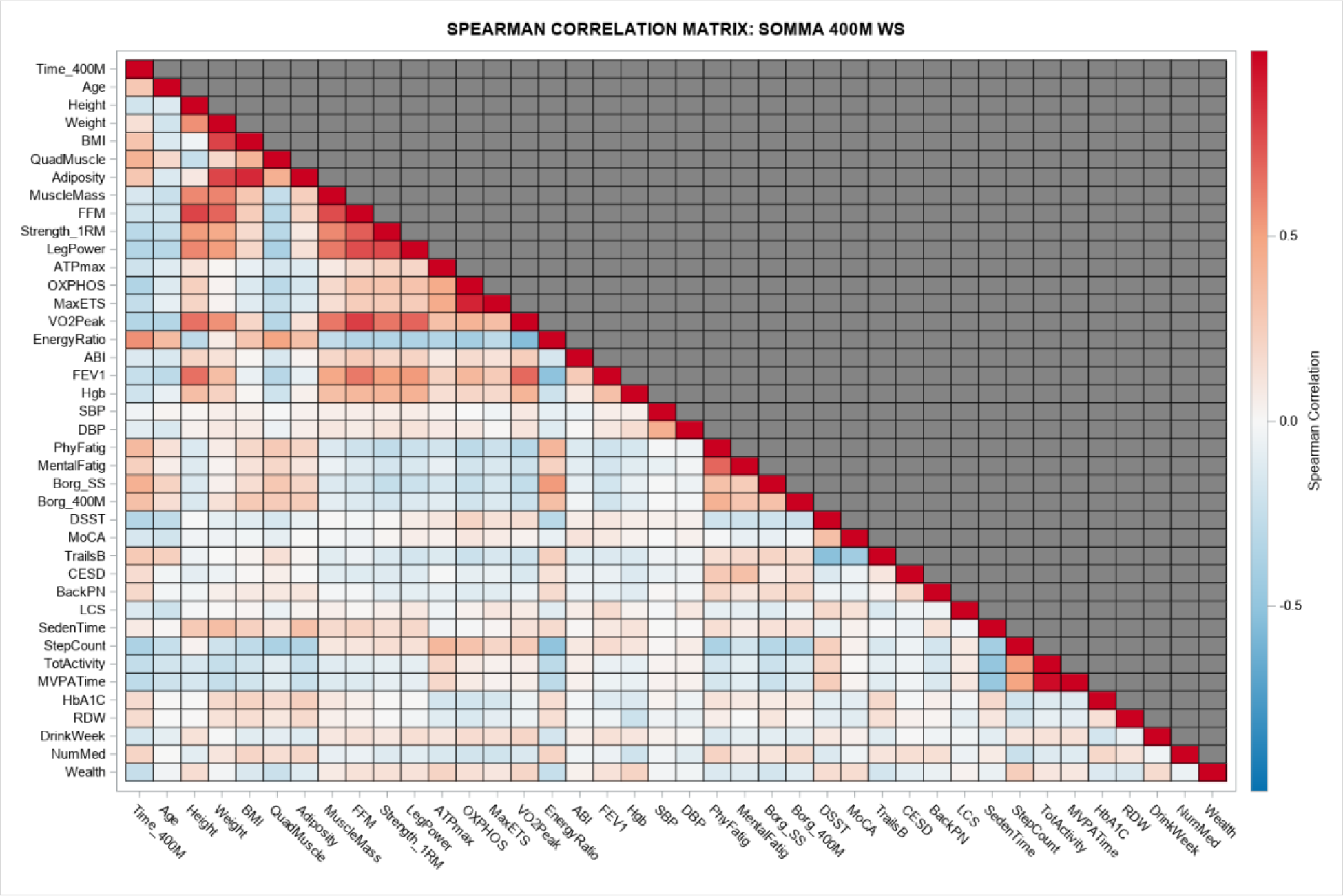
Heat map of correlations between potential determinants of 400m walking speed. *All bivariate correlations assessed here use variables expressed continuously, even if they were treated as ordinal variables in other analyses within this report.

**Supplemental Table S1.** Selected characteristics of the SOMMA cohort with number of missing.

| Characteristic | Number of missing |
| --- | --- |
| Sex | 0 |
| Race | 0 |
| Hispanic/Latino | 0 |
| Age | 0 |
| 400m walk time (sec) | 0 |
| Weight (kgs) | 0 |
| Body mass index (kg/m<sup>2</sup>) | 0 |
| Peak leg power (Watts) |  |
| Men | 13 |
| Women | 23 |
| Time in moderate to vigorous activity (min/d) | 56 |
| Sedentary time excluding in-bed time (min/d) | 194 |
| MoCA Total score | 14 |
| Anemia | 15 |
| History of chronic obstructive pulmonary disease | 3 |
| History of heart failure | 3 |
| History of stroke | 3 |
| History of diabetes | 3 |
| History of a diagnosis of depression | 4 |
| Current smoker | 5 |
| Number of drinks in the past 12 month | 14 |
| Ankle-arm blood pressure index (Lowest of Left and Right) < 1.0 | 69 |
| Peripheral neuropathy by monofilament test | 18 |
| Often/always hip or knee stiffness | 3 |
| Pittsburgh Fatigability Scale Physical score | 6 |
| Ratings of Perceived Exertion Fatigability at end of the steady speed test | 37 |
| VO<sub>2</sub>peak (volume of oxygen consumption) (mL/min) |  |
| Men | 17 |
| Women | 42 |
| Energy cost-capacity Ratio | 66 |
| Maximal mitochondrial oxidative phosphorylation (pmol/(s*mg)) | 134 |

**Supplemental Table S2.**
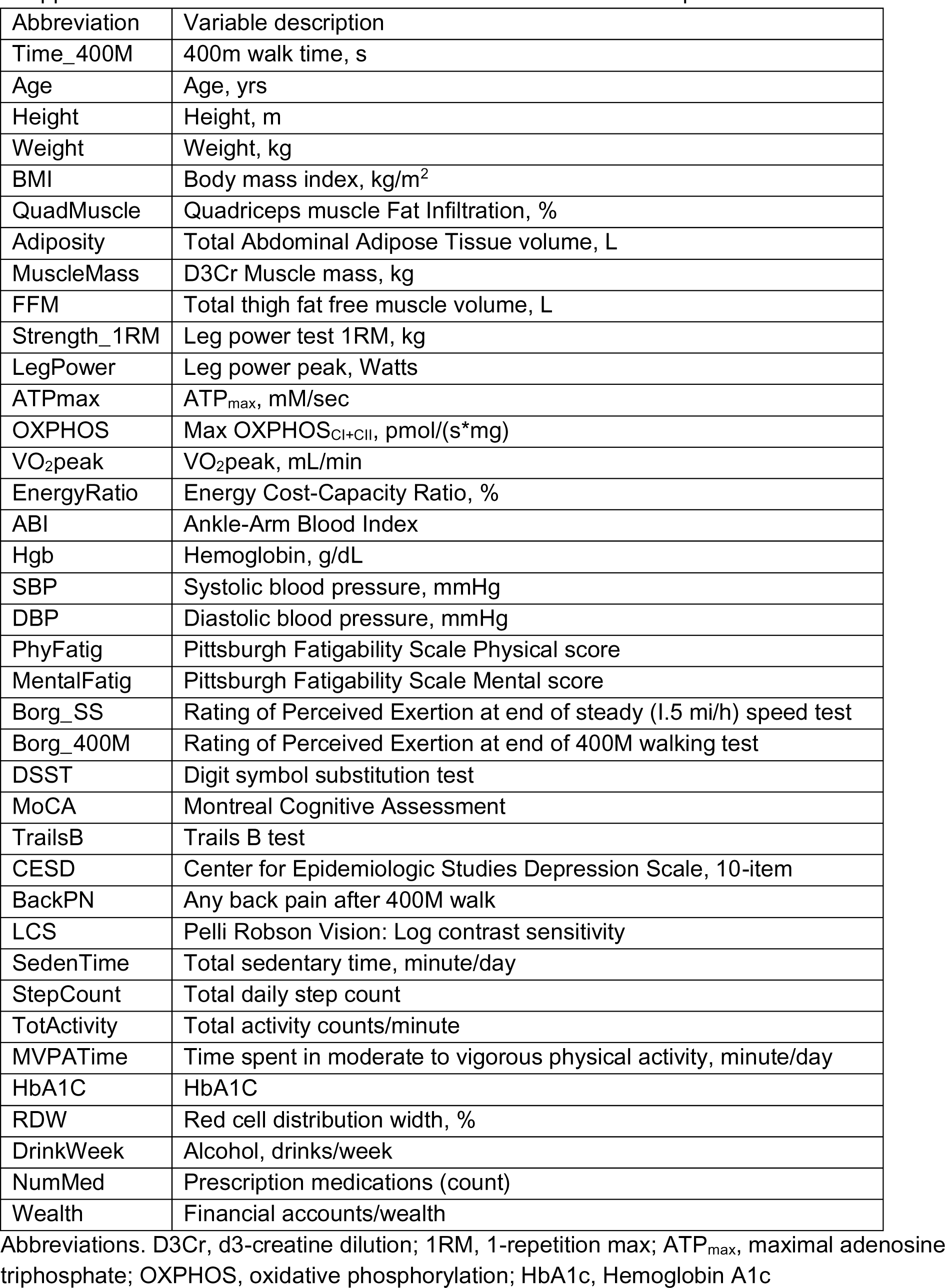
Abbreviations and definitions for the heat map.

